# Covid-19: Comparisons by Country and Implications for Future Pandemics

**DOI:** 10.1101/2021.05.29.21258056

**Authors:** Bangor Lewis Mehl-Madrona, Maine Orono, François Bricaire, Adrian Cuyugan, Jovan Barac, Asadullah Parvaiz, Ali Bin Jamil, Sajid Iqbal, Ryan Vally, Meryem Koliali, Mohamed Karim Sellier

## Abstract

**Background:** We set out in this paper to compare Covid-19 results by country to better understand the factors leading to the differing results found internationally.

**Methods:** We used publicly available large datasets to explore differences by country for Covid-19 mortality statistics. We continuously challenged our projections with reality and numbers from countries around the world, allowing us to refine our models and better understand the progression of the epidemic. All our predictions and findings were discussed and validated from a clinical viewpoint.

**Results:** While no lockdown resulted in higher mortality, the difference between strict lockdown and lax lockdown was not terribly different and favored lax lockdown. Only one of the top 44 countries had long and strict restrictions. Strict restrictions were more common in the worst performing countries in terms of Covid mortality. The United States had both the largest economic growth coupled with the largest rate of mortality. Those who did well economically, had lower mortality and less pressure on their population. Yet they had less mortality than average and less than their neighbors.

**Conclusions:** Countries with the least restrictions fared best economically. Some of them fared well in terms of mortality, even better than neighboring countries with similar social structures and more severe restrictions. The mortality rates in the USA, however, appeared to suffer from very high obesity rates. Norway and the northern European countries have less strict restrictions from the rest of Europe and had lower mortality rates. COVID-19 mortality was associated with vitamin D status.

## Introduction

The data needed to accurately track the transmission and impact of COVID-19 have been hard to collect. The available data do not provide the full picture of the epidemic, are not standardized among countries, and are not always standardized within regions of the same country. Understandably, most governments (and modelers) have focused their efforts on in-country tracking and predictions, making international comparisons difficult. Data are defined and collected differently from country to country, from period to period (and sometimes even within a country at the same period).

Despite these difficulties, mortality from COVID-19 is clearly higher in some countries than in others. Many factors could have a role in this disparity, including differences in proportion of elderly people in a population, general health, accessibility and quality of healthcare, and socioeconomic status. We set out in this paper to compare Covid-19 results by country to better understand the factors leading to the differing results found internationally.

## Methods

We formed a multi-disciplinary team that included an infectious diseases physician, data scientists, and software developers. We continuously challenged our projections with reality and numbers from countries around the world, allowing us to refine our models and better understand the progression of the epidemic. All our predictions and findings were discussed and validated from a clinical viewpoint. We used publicly available large datasets [1-11]

## Data Preparation and Analysis

Data was transformed to fit parametric distributions prior to statistical analyses, which were performed with Student’s t-test, linear regression and post-hoc tests. Especially for ordinary least squares regression, natural logarithmic transformations were done to remediate normality violations in the standardized residuals. Interpretation on the final model of the regression and analysis of variance were adjusted because of logarithmic transformations. For correlational studies, whether continuous or count data were used, both Pearson’s and Spearman’s rank correlational tests were done according to the data used. Normality assumptions were not made as we used non-parametric correlation tests to overcome nonlinearity. Most of these normality violations related to bimodal distribution, so running Pearson’s correlation tests provided a complete description of the association [12]. Also, power in significance testing were also done to ensure that for some data points that have very minimal number of samples, this was considered that the interpretation of significance is practically taken into consideration. For cross-correlation of stochastic processes proper diagnostics were done to ensure the absence of autocorrelation processes that could signal the delayed copy of itself from its own function of lag. Some weakpoints of the analysis that a limited area (i.e. UV Index, ozone and average daily ridership) were correlated to the national level counts or vice-versa with case counts, death counts, density GDP, etc with daily ridership. This is due to limited public data available. All significance levels are set at 0.05, otherwise specifically stated.

Data were collected at March 28, 2021 for Europe specific analyses and at November 30, 2020, for world comparison analyses.

### Excess mortality

was assessed using Euromomo charts, in which data points above the red line represent increases in excess mortality. Euromomo charts provide z-scores for excess deaths. This was cross-referenced with the Oxford dataset. The Oxford COVID-19 Government Response Tracker told us when each country imposed lockdowns and other restrictions. Spain imposed a lockdown on 14 March when they had 6300+ cases and almost 200 deaths. Italy imposed a targeted stay at home order on 23 February and a general population lockdown on 10 March when they had 10k+ cases and 630+ deaths. France imposed a lockdown on 17 March when they had 7.6k+ cases and 140+ deaths. The UK imposed a general population lockdown on 23 March when they had 12k+ cases and 360+ deaths. Greece imposed a lockdown on 23 March when they had 650+ cases. Malta imposed restrictions on gatherings on 10 March when they had 5 cases. Portugal imposed a lockdown on 19 March when they had almost 800 cases. Belgium imposed lockdown on 13 March when they had 559 cases and 3 deaths. Restrictions on large gatherings and public places were obtained from the Oxford data and from the Wiki France COVID timeline. Japan imposed a stay at home order on 8 April when they had 4400+ cases and almost 100 deaths (Wikipedia history of events). Tajikistan imposed a lockdown on 9 May when they had 610+ cases and 20 deaths, while Kazakstan imposed a strict lockdown on 19 March when they had 44 cases and no deaths yet. [April 1st (Week 14) on Euromomo was the highest peak in excess deaths. The Centre for the Mathematical Modeling of Infectious Diseases published estimated R0 figures and rates of growth and doubling time per country (https://dataverse.harvard.edu/dataverse/covid-rt).

### Environmental relationships

were tested using Pearson correlation analysis. The plots and values can be downloaded from https://peritusservices-my.sharepoint.com/:p:/g/personal/adrian_peritus-services_com/EeQWxQGX-s5PjfC5cbf2Bw8BHxFs9Njywh06FULKsyFyEA?e=XkPYn4. The relative humidity of cities was taken and aggregated into monthly average and quarterly average and then correlated with the national level counts of cases of deaths per country. This was found to be significantly correlated at 0.10 alpha (see link for the plot). UV Index and Ozone levels were taken from the TEMIS satellite dataset, which was cross correlated with the daily national level counts of cases and deaths per country (see https://public.tableau.com/profile/foxyreign#!/vizhome/UVIndexandOzone/Cross-Correlation). The cross-correlation dashboard based on selected European cities can be viewed at https://public.tableau.com/profile/foxyreign#!/vizhome/UVIndexandOzone/Cross-Correlation. This data does not adjust for average levels of melanin in each country. UV Index and Ozone Cross-Correlation data are found at https://public.tableau.com/profile/foxyreign#!/vizhome/UVIndexandOzone/Cross-Correlation.

We used Wikipedia pages of selected countries to obtain GINI Index, GDP per Capita, Density, Population count and Average Daily Ridership. We used Accuweather, Weather.com, and Weatherspark.com to obtain data on sunshine and relative humidity as monthly averages. Analysis was done with pair-wise Pearson product-moment correlations. We used 18 European cities, including Paris, Herault, Bouches du Rhone, Loire Atlantique, Nord, Brussels Capital Region, Community of Madrid, Barcelona, MilanRome, Porto Metro Area (North), Lisbon Metro Area, Copenhagen Urban, Stockholm County, Rio de Janeiro State, Sao Paolo State, Miami-Dade County, and New York City. Case counts, death counts, cases per capita and deaths per capita exhibited skewed distributions. Non-parametric methods were used.

Time-series with lag regression was used to compute the **mean incubation period** for change in percentage of deaths and not the absolute count of deaths. The screenshot of the model summaries are available at https://peritusservices-my.sharepoint.com/:p:/g/personal/adrian_peritus-services_com/EQhDar3AuGZAhRmUvvrKmyABpfHbSGJ8xdkMs7vnKg0K7Q?e=1K3KHy. Pre-selection of the lag to be included in the lag regression was done using the cross-correlation but the time-series did not undergo pre-whitening nor was it adjusted for seasonality.

### Data sources included the

UV station data based on TEMIS satellite ozone data, available at https://www.temis.nl/uvradiation/UVarchive/stations_uv.php (Cloud-free erythemal UV Index and local solar noon ozone values, date range from 1 Mar to 25 May 2020), COVID-19 Coronavirus Pandemic, Worldometers, date range from 1 Mar to 25 May 2020, and COVID-19 Community Mobility Reports, Google, date range from 1 Mar to 25 May 2020 - https://www.gstatic.com/covid19/mobility/Global_Mobility_Report.csv.Selected cities were joined that matched between Google Mobility and TEMIS Satellite datasets. There were 7 cities included Venice, Paris, Madrid, Copenhagen, Bern, Stockholm, and Lisbon. The daily death change percentage has extreme values so any death change percentage above 200% was replaced, setting the limit to 200% (i.e. the maximum was 3,600% Sweden from 2 to 80+ deaths in just two days!). We ran the 1^st^ iteration of the model using the computed for cross-correlation of available data and picked out the top 3 correlation coefficient values under 0.05 significance:

- UV Index – lag +7 and +11
- Transit – lag +11, +17, +24
- Residential – lag 10, 17, 24

Then we ran the 2^nd^ iteration of the model using the computed for cross-correlation of available data and picked out the highest correlation coefficient values under 0.05 significance:

- UV Index is significant at 0.05
- Transit is not significant
- Residential is significant at 0.10

Model residuals were tested for normality. We ran final iteration of the model with the above significant independent variables with lag. Model residuals are normal.

Data sources for sunlight included the COVID-19 Coronavirus Pandemic, Worldometer, (accessed on 5 May 2020 at https://www.worldometers.info/coronavirus/), Selected cities by sunshine duration, Wikipedia (accessed on 5 May 2020 at https://en.wikipedia.org/wiki/List_of_cities_by_sunshine_duration, and Selected cities by average temperature, Wikipedia (accessed on 5 May 2020 at https://en.wikipedia.org/wiki/List_of_cities_by_average_temperature). We used Pearson product-moment correlation and scatterplot visualization.

### Average daily ridership

of some cities was correlated with counts of cases and deaths where the metro was located using the Google Mobility Cross-Correlation dashboard. Transit stations had a positive correlation at +25 days lag with death count on the following locations, significant at p < 0.05 for France (0.26), New York State (0.367), and Italy (0.406). Demographics, daily ridership levels, and some national economic scores were used in correlation analysis. The plots may be viewed at https://peritusservices-my.sharepoint.com/:p:/g/personal/adrian_peritus-services_com/EU8Skd0VqvdNmgSZ_Q-x-YABmc3T2WMcACAuW_esvUU3gg?e=3asFda. Correlation coefficients for mobility data were derived from cross-correlation analysis done on each variable of the Google mobility dataset with the COVID cases and deaths and can be found at https://public.tableau.com/profile/foxyreign#!/vizhome/GlobalMobilityReport_15917747268540/GoogleMobilityReportDeaths-A4size.

The technical specifications and software used to calculate all **regression models** are available at https://peritusservices-my.sharepoint.com/:w:/g/personal/adrian_peritus-services_com/EQGtrtuLajBIoqxUacfwVCUB4XEPH31thzXCycV6eLQA_A?e=IZuYlr. Model selection was either based on adjusted R^2^ or f-statistic p-value using the typical Gaussian linear regression that assumes normal distribution. Log-log normal distribution was used to compute the change in deaths per million with percentage of obesity. Further information is available at https://peritusservices-my.sharepoint.com/:i:/g/personal/adrian_peritus-services_com/Eaj5eFSTyfBKtPV-YfWPAJIBLXA3FEqDgWUFQZbi3tW3Zg?e=y97F3M.

The demand for air conditioning of certain countries from 2018 and 2019 was correlated with cases and deaths. See screenshot: https://peritusservices-my.sharepoint.com/:i:/g/personal/adrian_peritus-services_com/EQZr7N-YIlpDhVogS24e0VQBf0ZsqlyjCjZPkzTqHJR3Cw?e=agsm4s. Data sources included https://www.jraia.or.jp/english/World_AC_Demand.pdf and the Cumulative COVID-19 Our World in Data cases, which showed numbers of deaths and number recovered as of 24 June 2020. We used Pearson product-moment correlation.

### Obesity

We used Our World in Data, cumulative count as of 4 Jul 2020 - https://github.com/owid/covid-19-data/blob/master/public/data/owid-covid-data.csv and the Central Intelligence Agency of obesity adult prevalence rate (Country Comparison:: Obesity - Adult Prevalence Rate, 2016) - https://www.cia.gov/the-world-factbook/field/obesity-adult-prevalence-rate/country-comparison. Countries were classified by continent according to their geographic location. Total (cumulative as of 4 Jul 2020) COVID deaths per million dependent variables was transformed using natural logarithmic + 1 to eliminate infinity values. We ran a log-normal ordinary least squares regression model using effects coding (sum-to-zero contrast, setting Africa as the reference category) with the following independent variables:

- Obesity prevalence rate nested with the obesity per continent
- Population density
- Aged 70 older
- GDP per capita
- COVID death rate
- Diabetes prevalence
- Female smokers
- Male smokers
- Hospital beds per thousand
- Life expectancy

We ran normality test on the model residuals and checked generalized variance inflation factors for multicollinearity. We ran ANOVA to check the difference in the means and then ran Tukey post-hoc tests to check for pair-wise difference in means. The model coefficients were transformed back using exponents to interpret the model values using Y percent in increase/decrease = [(exp(beta coefficient – 1) * 100].

### Face Mask Data

We used the COVID Tracking Project, accessed 8 Aug 2020, at https://covidtracking.com/data. Facemask use during the COVID-19 pandemic in the United States was accessed 8 Aug 2020 using https://en.wikipedia.org/wiki/Face_masks_during_the_COVID-19_pandemic_in_the_United_States. The COVID tracking project is a time-series dataset that has different variables with daily counts for each state. This dataset was joined with the Wikipedia dataset that classifies the dates if the counts occurred during ‘pre-mask’, ‘post-mask’ or ‘no law’. We used Quassi-poisson regression, zero-inflated Poisson regression, and auto-regressive time-series regression, plus median analysis with random sampling bootstrapped and 5,000 replications of pre-mask vs post-mask and no law.

### Patient and Public Involvement

The research questions and outcome measures were those being discussed widely on all news channels. Given that we were all potential patients, we were all involved in the design of the study in that we asked the questions that potential patients were asking in the media.

## Results

### Country Comparisons

In Figure 1, we present data from Eurostats on excess mortality per country comparing Jan 2020 – Jan 2021 to 2016-2020 periods. We observe that the Scandinavian countries have the least excess mortality. We have selected countries to discuss that encompass the range from minimal to high.

**Figure 1.**
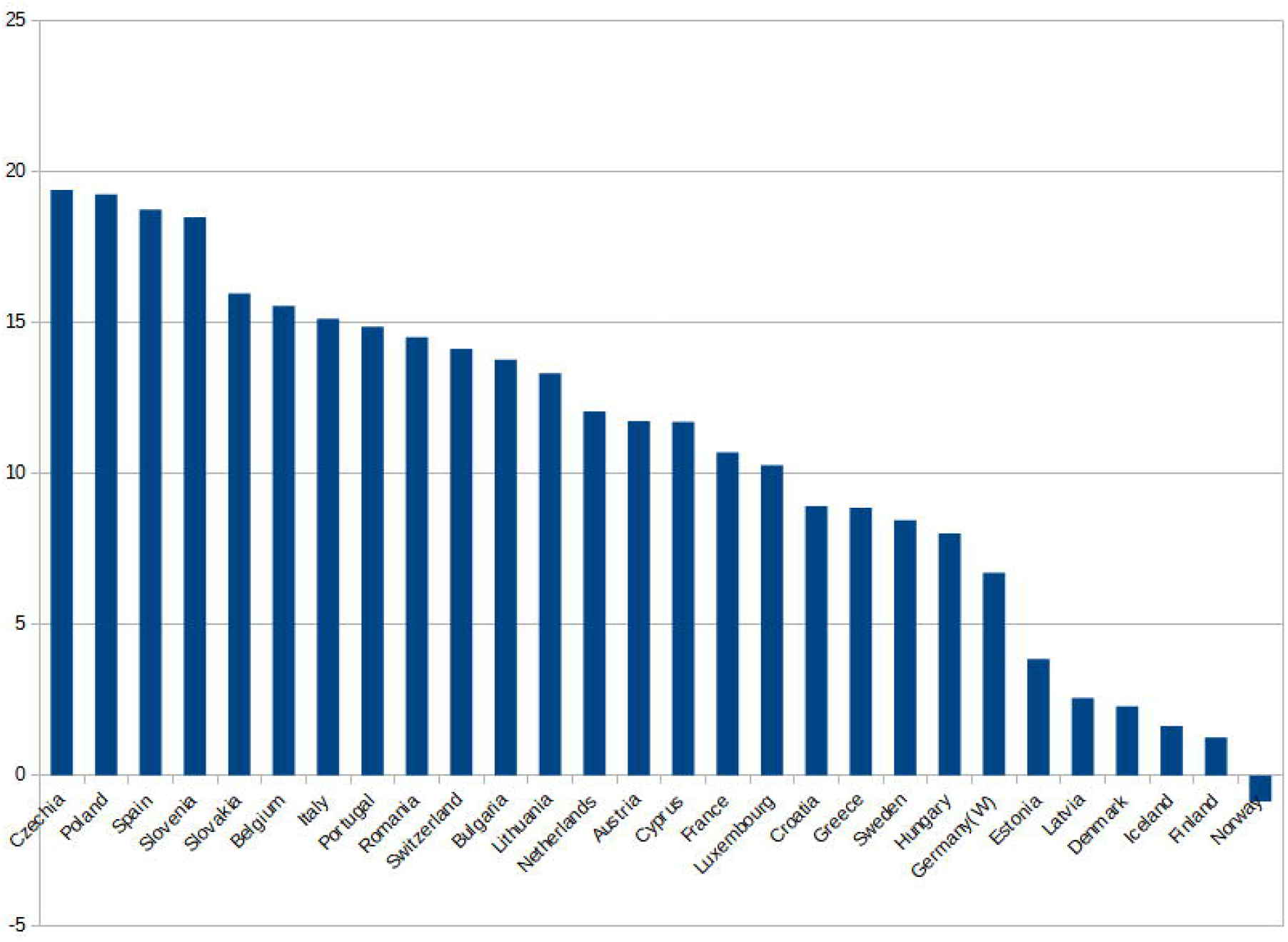
Comparison of excess mortality by country.

#### Digging into epidemic phases by country

##### Sweden

Sweden allowed the virus to follow much of its natural course due to the limited restrictions imposed by the government and to its northern lattitude and reduced sun exposure. The actual number of infected cases of Covid-19 can only be extrapolated such that all Infection Fatality Rate (IFR) values are approximations since the true number of infections is unknown. If we account for specific antibodies detected through serology tests and add memory immunity T cells, cross immunity, and cellular immunity, we find values between 14% and 30%, which puts us in a range between 1,400,000 and 3,000,000 cases. This results in an IFR or lethality between 0.17% and 0.34%.

We note a 3,725 count in excess mortality instead of the 5,420 Covid declared deaths. Some of the extra deaths could be due to those from comorbidities that were attributed to Covid. IFR may actually be lower and would drop to a range between 0.12 % to 0.34 %.

Ledberg (2020) explored the epidemic, only looking at the aggressive window from the end of March to early May reaching a count of 4,857, representing a 10.5 % excess mortality, lower than the 1940/1941 influenza outbreak in terms of mortality/capita. Looking at a wider window gives a better perspective as to actual excess mortality. Covid’s effect on a naive population has led to a maximum increased mortality of 8 % compared to influenza. in previous years, assuming no deaths were caused by influenza, the effect of Covid on the population in Sweden where little restrictions were applied was an 8% increase in death compared to previous years, assuming all excessive death was caused by Covid and none by Influenza.

The 2020 mortality from all causes was 0.48 %. Within that the excess mortality related to Covid was 0.04%. This is no comfort for those who lost loved ones, but as immunity builds, future eventual outbreaks are likely to be milder and more can be done to prevent mortality. Over a semester of Covid in Sweden, where the population was recommended to be cautious and large events were cancelled, but also where influenza seemed almost absent, the excess mortality over a semester was 8 % to 10 % more severe than that of previous years in which there were influenza outbreaks.

Sweden is a country that does not benefit from sun, and where, like much of Europe, there is significant obesity. On the other hand, Stockholm, the main city, is less dense than some European capitals.

A the time of writing this paper late March, 2021, Sweden has not been witnessing excess mortality for 6 consecutive weeks.

##### Denmark

In Denmark, the epidemic effect on mortality is invisible. Over the first half of 2020 an under-mortality can be observed despite a short lockdown of places. We observed a difference between under-mortality and Covid death suggesting a possible Covid death attribution that may be attributed to comorbidities. Excess mortality due to Covid-19 was -0.01 %

During the fall of 2020 and the winter of 2021, Denmark did witness a moderate excess mortality but still seems to be spared.

##### Belgium

During phase 1 of the epidemic between February and May 2020, Belgium was one of the 3 hardest hit countries in the world despite a severe prolonged lockdown. It declared 9,776 Covid deaths. Excess mortality was 3,993, confirming the same excess in Covid counts with respect to excess mortality as observed for Sweden and Denmark. This difference could be caused by two possible explanations each of which could contribute to this 60 % overcount. Either the Covid situation spared up to 5,783 lives or simply 5,783 deaths occurred with patients who died of comorbidities in the presence of Covid, but actually died of their comorbidities. Belgium has a population of approximately 11,486,000. Excess mortality in 2020 was around 7% for the top 3 hardest hit countries during initial epidemic phase. A second phase started in October, 2020, leading again to visible excess mortality until mid-December. As per Be-MOMO, since mid-December, there was no notable excess mortality in Belgium. Since February, 2021, in spite of some mortality attributed to Covid, mortality seems to be lower than normally expected.

At the time of writing this paper late March, 2021, Belgium has not observed any excess mortality for 6 consecutive weeks.

##### France

France Insee’s data provides a similar pattern to that of Sweden with an excess mortality of 6.73 %, corresponding to 17,691 people. This is lower than the 29,779 Covid deaths declared by France. If we set aside excess mortality caused by influenza or lockdown and assume that all of that excess mortality is attributed to Covid-19, that figure is still lower by 40 % than the COVID-19 death total, indicating most likely a large comorbidities factor attributed to COVID. Given that May and June 2020 showed under-mortality, the excess mortality displayed during the Covid episode is comparable to that of the 2016 – 2017 influenza outbreak.

When we examine the recent severe Influenza epidemic in 2016-2017 over the months of December 2016 to February 2017 and compare mortality to that of February 2020 to April 2020, the Covid year shows an excess mortality of 1.42 % in one of the 10 hardest countries in the World as per death per million inhabitants. The under-mortalities that followed further confirm the role of comorbidities in Covid death counts which could turn out to be 30 % to 60 % of Covid attributed deaths. Having a longer look at history gives us a better insight into what happened in one of the hardest hit countries in the world, but also one that displayed sufficient data transparency because of its structures. In France, 2020 in terms of mortality was slightly milder, or comparable to 2016/2017, 1973, 1997, and 2000 influenza outbreaks and milder than 1969, 1956, 1963, and 1962.

Monthly Death (INSEE data)/Population (Estimate of linear evolution from 42 Million to 67 Million). This suggests a disproportion between the epidemic’s impact and the populations’ overall reactions which could lead to actions and regulations that are counter-productive on the physical, physiological, and the psychological health of populations, rendering them more fragile against Covid-19.

Ined’s recent report indicates for all 2020, we see an excess mortality of 42,000 taking into account population increase and aging, whereas Covid attributed Mortality was slightly below 68,000. That same report finds that Covid attributed deaths follow similar proportions to mortality amongst different age groups further confirming the role of comorbidities and the importance of a population’s general health.

France suffered 2 milder successive phases compared to its initial phase.

At the time of writing this paper, France is still suffering from a moderate excess mortality that maybe subsiding in April as immunity levels rise and weather becomes more favourable.

###### Portugal, Slovenia, Poland, Czechia, Slovakia, Lituania, Greece, Croatia

were spared during the initial phase possibly due to travel restrictions and spring arrival before the outbreak only to suffer a severe outbreak in late fall, 2020, and winter 2021. At that stage, all European countries had adopted community mitigation strategies. These countries are not currently suffering from significant excess mortality.

###### Latvia and Estonia

**s**eems to have faced much of its outbreak in February and March of 2020 and are now catching up with Sweden.

###### Finland, Iceland and Norway

seem to have skipped their turn with moderate or no excess mortality.

### Country Comparisons by Strictness of Lockdown

When we compare countries with very different lockdown policies, we see an average of 0.61% of losses of population in Italy, France, Spain, compared to more lax lockdown policies of Denmark and Austria (0.05%), and compared with no lockdown policies (Sweden) which showed 0.795% losses of population. While no lockdown resulted in higher mortality, the difference between strict lockdown and lax lock-down was not terribly different and favored lax lockdown during 1^ST^ epidemic phase. During second epidemic phase, Sweden witnessed a negligeable loss of population whereas France, Italy and Spain still witnessed significant mortality. Norway, Finland, and Denmark did not witness any significant excess mortality.

### Covid Mortality by Country: a broader look beyond Europe

Table 1 presents the best 44 performing countries in terms of mortality and the worst 44 countries going from number 104 to number 151 through November, 2020. Only one of the top 44 countries had long and strict restrictions. Strict restrictions were more common in the worst performing countries.

**Table 1.**
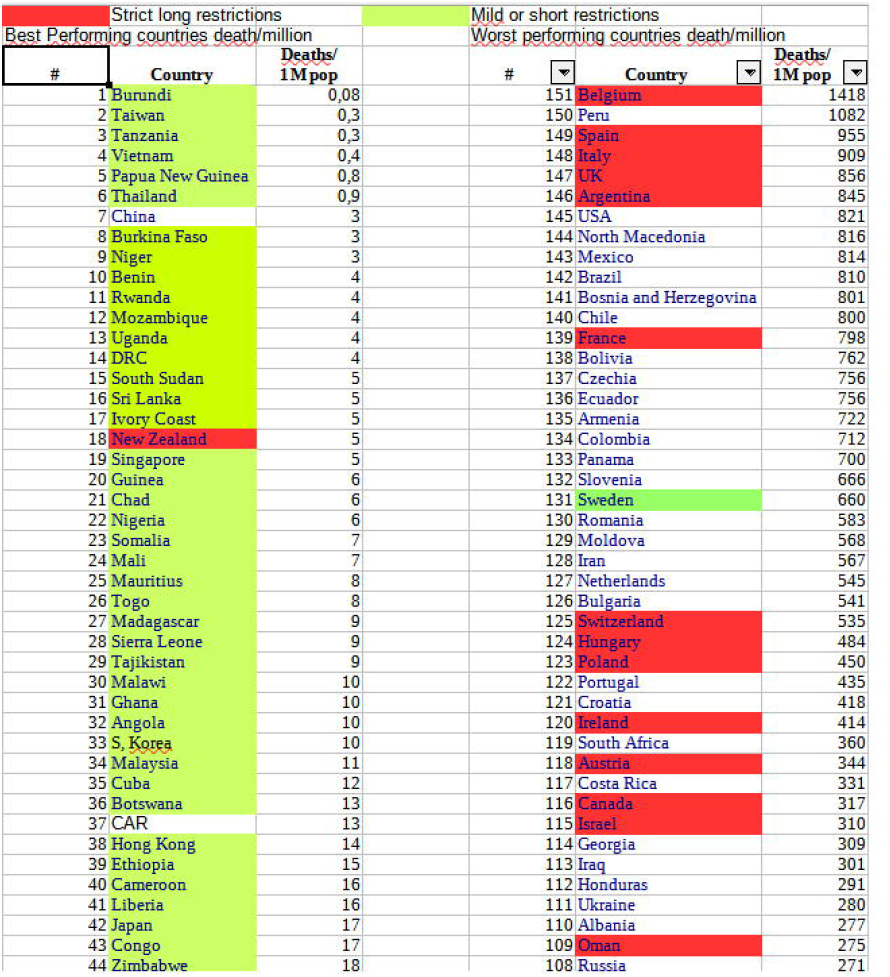
Rankings by country in terms of Covid-19 mortality rates in relation to strictness of restrictions and lockdown policy.

| Strict long restrictions |  |  | Mild or short restrictions |  |  |
| --- | --- | --- | --- | --- | --- |
| Best Performing countries death/million |  |  | Worst performing countries death/million |  |  |
| # | Country | Deaths/<br>1M pop | # | Country | Deaths/<br>1M pop |
| 1 | Burundi | 0,08 | 151 | Belgium | 1418 |
| 2 | Taiwan | 0,3 | 150 | Peru | 1082 |
| 3 | Tanzania | 0,3 | 149 | Spain | 955 |
| 4 | Vietnam | 0,4 | 148 | Italy | 909 |
| 5 | Papua New Guinea | 0,8 | 147 | UK | 856 |
| 6 | Thailand | 0,9 | 146 | Argentina | 845 |
| 7 | China | 3 | 145 | USA | 821 |
| 8 | Burkina Faso | 3 | 144 | North Macedonia | 816 |
| 9 | Niger | 3 | 143 | Mexico | 814 |
| 10 | Benin | 4 | 142 | Brazil | 810 |
| 11 | Rwanda | 4 | 141 | Bosnia and Herzegovina | 801 |
| 12 | Mozambique | 4 | 140 | Chile | 800 |
| 13 | Uganda | 4 | 139 | France | 798 |
| 14 | DRC | 4 | 138 | Bolivia | 762 |
| 15 | South Sudan | 5 | 137 | Czechia | 756 |
| 16 | Sri Lanka | 5 | 136 | Ecuador | 756 |
| 17 | Ivory Coast | 5 | 135 | Armenia | 722 |
| 18 | New Zealand | 5 | 134 | Colombia | 712 |
| 19 | Singapore | 5 | 133 | Panama | 700 |
| 20 | Guinea | 6 | 132 | Slovenia | 666 |
| 21 | Chad | 6 | 131 | Sweden | 660 |
| 22 | Nigeria | 6 | 130 | Romania | 583 |
| 23 | Somalia | 7 | 129 | Moldova | 568 |
| 24 | Mali | 7 | 128 | Iran | 567 |
| 25 | Mauritius | 8 | 127 | Netherlands | 545 |
| 26 | Togo | 8 | 126 | Bulgaria | 541 |
| 27 | Madagascar | 9 | 125 | Switzerland | 535 |
| 28 | Sierra Leone | 9 | 124 | Hungary | 484 |
| 29 | Tajikistan | 9 | 123 | Poland | 450 |
| 30 | Malawi | 10 | 122 | Portugal | 435 |
| 31 | Ghana | 10 | 121 | Croatia | 418 |
| 32 | Angola | 10 | 120 | Ireland | 414 |
| 33 | S. Korea | 10 | 119 | South Africa | 360 |
| 34 | Malaysia | 11 | 118 | Austria | 344 |
| 35 | Cuba | 12 | 117 | Costa Rica | 331 |
| 36 | Botswana | 13 | 116 | Canada | 317 |
| 37 | CAR | 13 | 115 | Israel | 310 |
| 38 | Hong Kong | 14 | 114 | Georgia | 309 |
| 39 | Ethiopia | 15 | 113 | Iraq | 301 |
| 40 | Cameroon | 16 | 112 | Honduras | 291 |
| 41 | Liberia | 16 | 111 | Ukraine | 280 |
| 42 | Japan | 17 | 110 | Albania | 277 |
| 43 | Congo | 17 | 109 | Oman | 275 |
| 44 | Zimbabwe | 18 | 108 | Russia | 271 |

Table 2 shows similar data for the individual states in the United States. Three of the top 10 performers in terms of deaths/million had tough restrictions. Five of the 10 worst performers had tough restrictions.

**Table 2.**
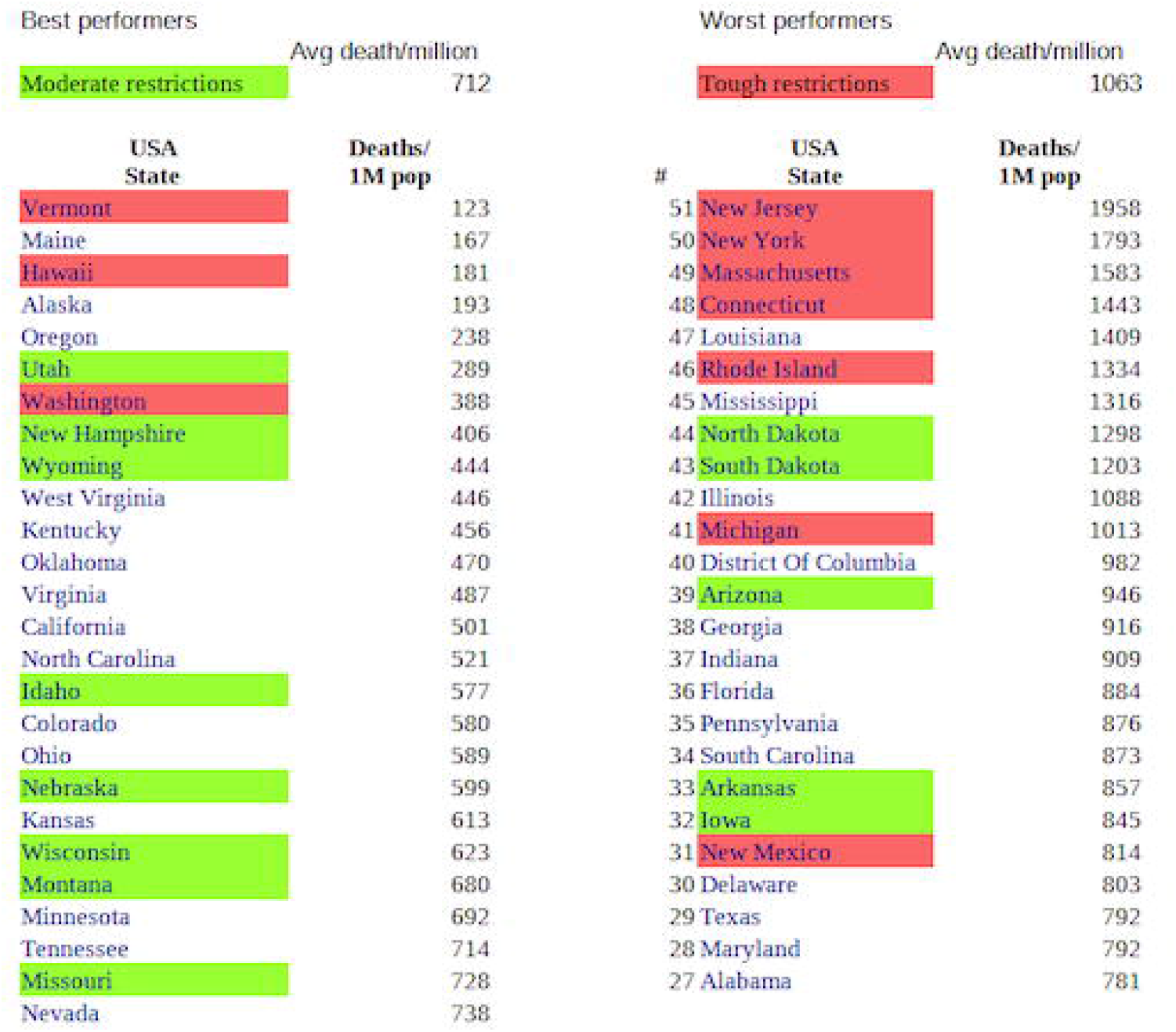
Rankings by State in the USA in terms of Covid-19 mortality rates in relation to strictness of restrictions and lockdown policy.

When we compare the mortality in deaths/million from weeks 1 to 41 for the years 2017, 2018, 2019, and 2020, we can visualize the actual increase in the death rate in 2020 when coronavirus appears compared to prior years. The year 2020 shows approximately 750 deaths/million more than what was found in previous years, which approximates the reported numbers of coronavirus-related deaths for the first 41 weeks of 2020, which was 247,500 excess deaths in the United States. The number of deaths/million in 2017 in the first 41 weeks of 2017 were 6,758; in 2018, 6,814; in 2019, 6,792; and in 2020, 7,551.

Table 3 shows that economic growth was possible in some countries despite large increases in mortality. The United States had both large economic loss and the largest rates of mortality. In general, countries with the least restrictions fared best economically. Surprisingly, some countries fared better with less restrictions than neighboring countries with similar social structures and stricter restrictions. These include Uzbekistan compared to Tajikistan, Morocco compared to Egypt, and Belarus compared to Poland and Lithuania. Also, surprisingly, some countries with relatively poor health care capabilities and little capacity to impose restrictions fared well. Guyana’s economic improvement was related to discoveries of large oil resources. Benin promoted construction projects to increase their gross domestic product (GDP). The GDP of the United States and Ireland benefited from digital and pharmaceutical sectors. Norway and other northern European countries fared better than other European countries. Their digital economies and oil exports benefited their economy. Egypt redirected its activities from tourism to building infrastructure. Egypt had a curfew, closed restaurants, instructed sick people to stay home, and advised people to eat fruits and vegetables. Vietnam quickly isolated infected clusters. Table 3 relates annual GDP growth (or shrinkage) compared to Covid-19 mortality rates.

**Table 3.**
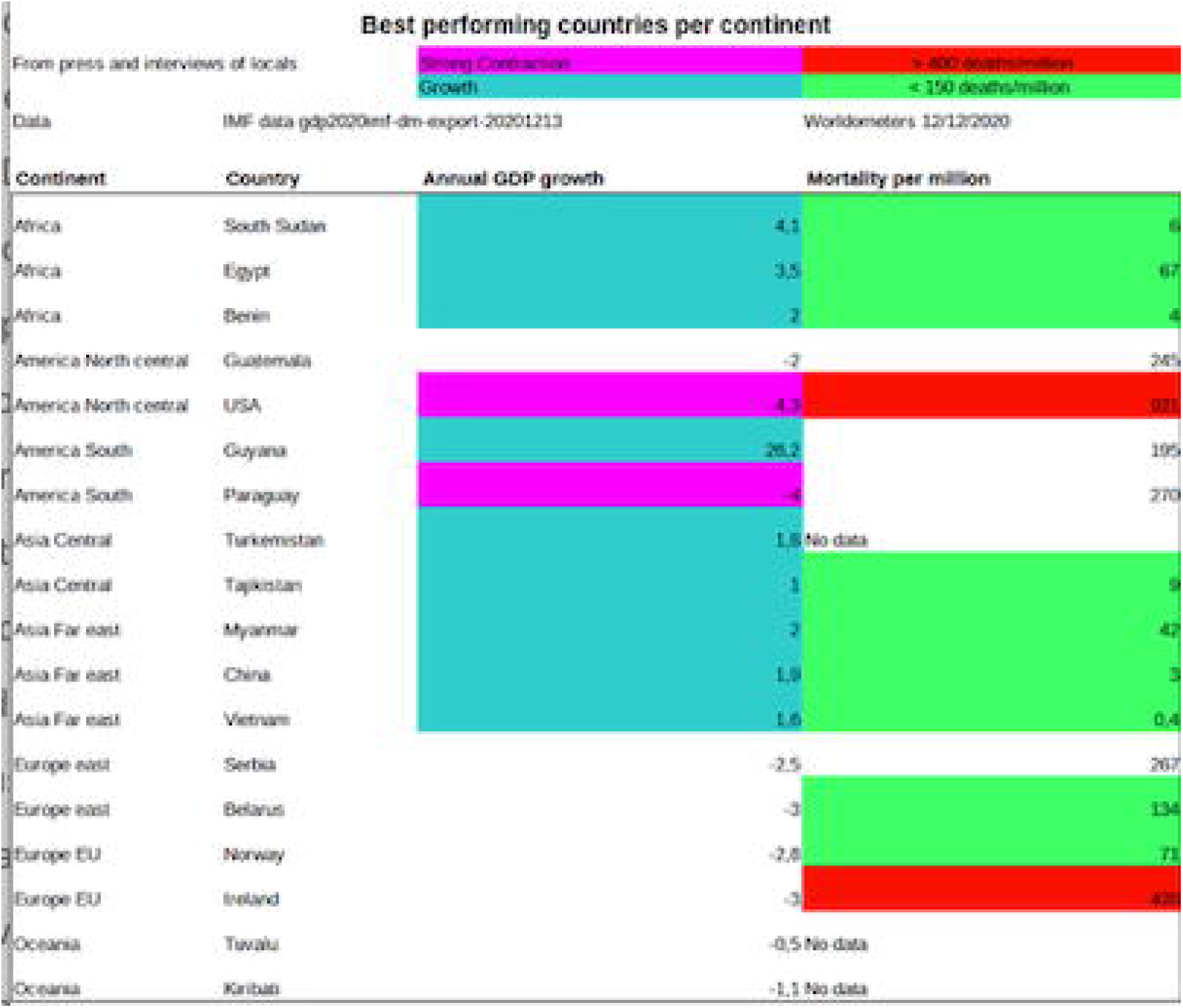
Best performing countries by continent in terms of gross domestic product and Covid-19 mortality.

Figure 2 shows that larger Covid-19 death rates are associated with larger losses in GDP, which is logical.

**Figure 2.**
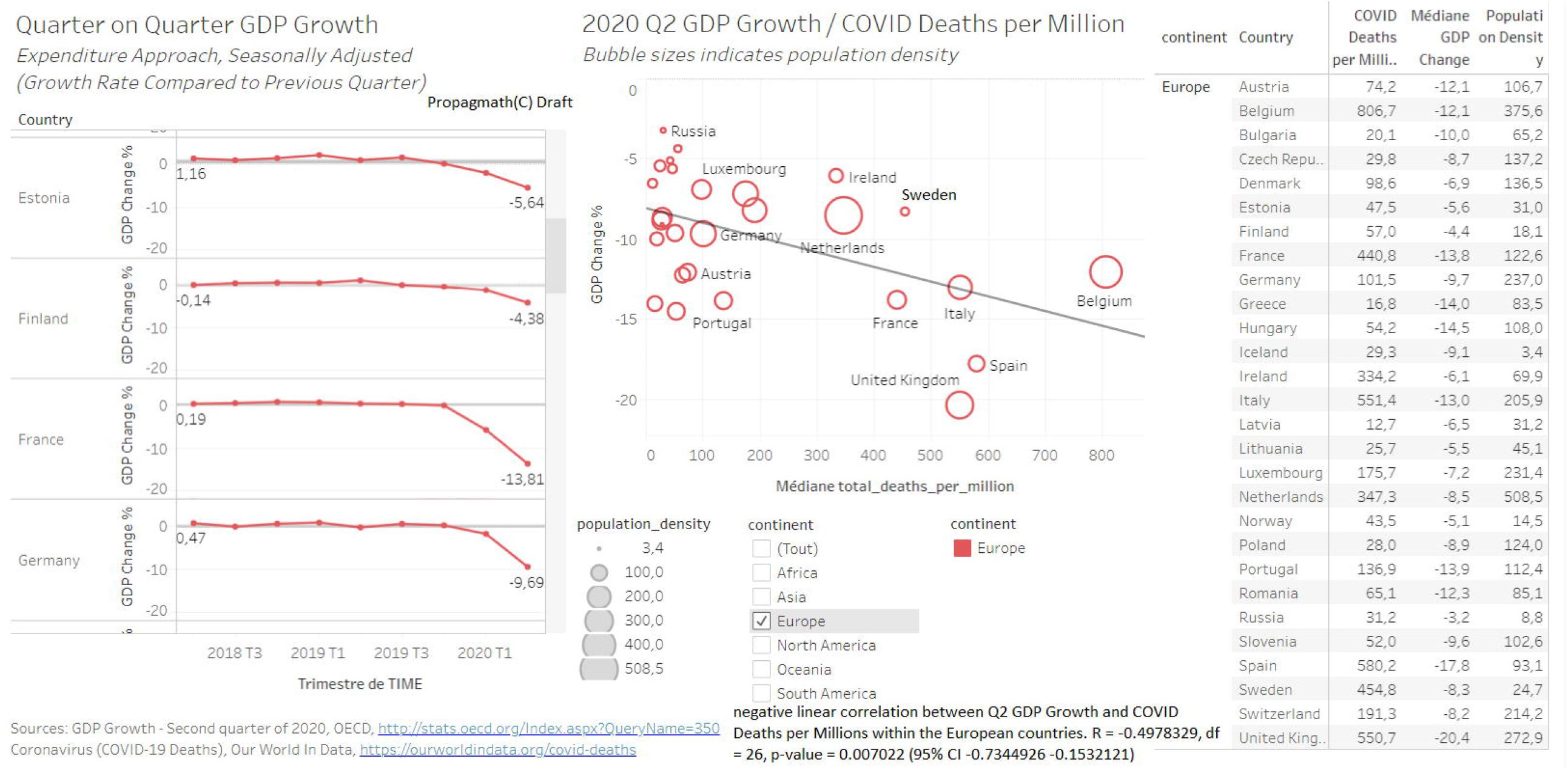
Correlation of change in gross domestic product and Covid-19 mortality rates.

Figure 3 presents the best performing countries economically compared to their Covid-19 death rate. As seen with correlation maps those who did well economically, had lower mortality and less pressure on their population. Yet they had less mortality than average and less than their neighbors.

**Figure 3.**
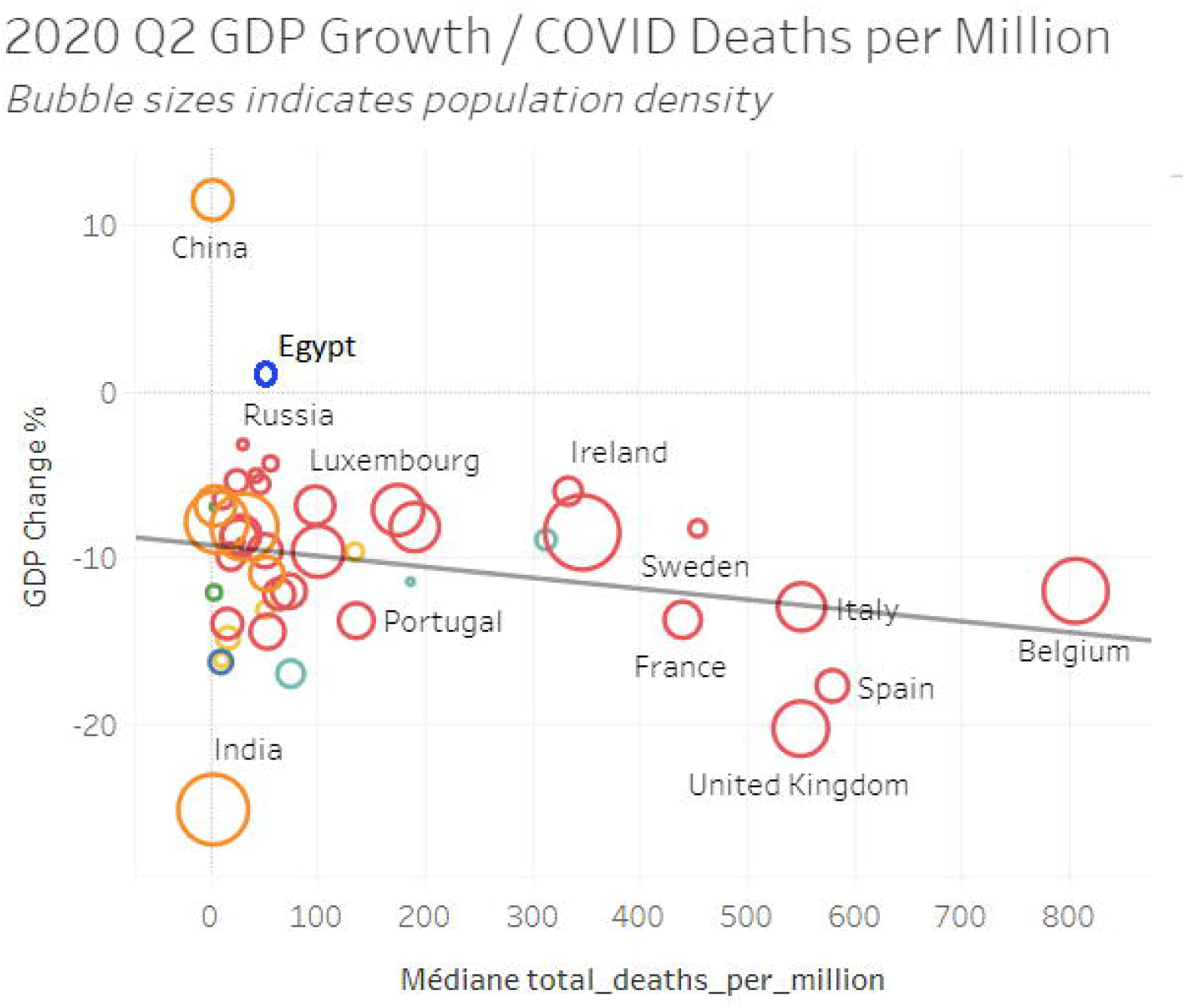
Change in gross domestic product and Covid-19 mortality rates in relation to population density.

### Sensitivity and resistance

Another significant factor in contagion is population resistance --the presence of a population whom, under normal circumstances would not get infected because of past immunity or other reasons as observed in the past. A healthy population with little obesity, good diet, and adequate levels of vitamin D would be more resilient than an unhealthy population. Children are less likely to contract the virus and less likely to transmit it, so the age of the population matters. For example, in the case study of the French aircraft carrier Charles De Gaulle, in a population of 1,760 who were sharing common dorms, common corridors, and common cantines for a month, 1,043 (59%) were infected, but the other 41% tested negative despite having been exposed to significant viral loads repeatedly. This is mostly a male and rather young population. It is possible this resistance may be lower in an older population and potentially higher in women and children. It is also possible that a higher fraction of the adult population may be resistant to the virus under normal exposure due to genetics, cross-immunity, past immunity or other factors. SARS-CoV-2 reactive CD4+ T cells were detected in 40%–60% of unexposed individuals, suggesting cross-reactive T cell recognition between circulating “common cold” coronaviruses and SARS-CoV-2 [18].

Recent large gatherings across the world in countries in open-air spaces with few masks did not lead to cluster formation in spite of massive testing further confirming that contamination happens indoors.

Japan, the oldest country in the world with some of the highest density cities and hardly any restrictions was not confronted to any signficant excess mortality. Japan has a superior diet, its elderly do not suffer from vitamin D defficiency, and it has virtually no obesity like much of Asia.

Practically all the large cities strongly affected during phase 1 and for which we have data had a milder phase 2. This is a sign of a possible start to build up sufficient mass immunity and protection to slow the epidemic in these cities. The reverse was true for many cities spared in phase 1 but heavily affected in phase 2 despite identical rules. The observation from London shows that the variant slightly enhances the need for mass immunity; the plateau of the curve in London shows that this complement would have occurred after 2 weeks of circulation. Yet, New York, Belgium, and Madrid were unaffected by the British variant possibly thanks to a higher level of natural immunity acquired at the time the variant arrived. Some cities seem less sensitive to the epidemic; if it is density or urban planning or composition of the population or another permanent fact remains to be determined.

## Discussion

On a year-to-year analysis, in spite of community mitigation strategies, except for a few countries far north, most countries seemed to catch up and these non-pharmacological interventions do not seem to be driving the epidemic. Their effect is modest compared to population health, urban structure, tourism roaming, obesity, pre-existing cross immunity, vitamin d levels, and weather. [Mehl-Madrona et al.]

Overall, most countries were hit in phases with a comparable outcome in spite of community mitigation strategies. Travel restrictions may have spared some regions in March 2021 only to catch up between October 2020 and March 2021.

Norway, Iceland, and Finland seemed to have been successful with targeted, limited community mitigation strategies. They never issued stay home mandates, used masks in dense closed places and kept much of the schooling system open. Their other limitations were short in time. It is possible that by targeting measures for a short time they had better adherence. It is also possible that vitamin D supplementation policies helped as well as low density lifestyle. One may also consider that these are relatively isolated countries with limited roaming, tourism, and international travel. Beyond Norway and Finland, lax lockdown policies could lead to people spending more time outdoors and receiving more UV radiation which could have improved vitamin D status.

Looking at a broader perspective, there is no indication that strict long measures reduced mortality. The data suggests the opposite.

Countries with the least restrictions fared best economically. Surprisingly, some of them also fared well in terms of mortality, even better than neighboring countries with similar social structures and more severe restrictions. Developing countries with little healthcare capabilities and limited ability to enforce restrictions tended to fare well. Countries with treatments, independent of the type of treatment, fared well. Does this mean that a placebo effect was in operation or is any treatment better than no treatment?

The mortality rates in the USA, however, may have suffered from very high obesity rates. Norway and the northern European countries have less strict restrictions from the rest of Europe and, also, had lower mortality rates. Was this from the longer summer days and earlier cold winter? Egypt fared well despite dense cities, relatively high obesity rates, and high co-morbidities among its older population (though on average its population is relatively young). Viet Nam fared well by isolating infected clusters in Da Nang and in hospitals. Turkministan chose to completely ignore the epidemic. Like similar countries with low obesity, low density, and a young population, they fared well. Most locations with a substantial first wave mortality and a milder second wave. Extended, prolonged restrictions did not necessarily reduce mortality but did adversely affect economics. Further investigation is needed to tease out the reasons for the wide variations in countries.

Others have pointed to the variations in vitamin D status by country in relation to Covid-19 outcomes. Mitchell [19] notes that SARS-CoV-2, the virus responsible for COVID-19, emerged and started its spread in the Northern hemisphere at the end of 2019 (winter), when levels of 25-hydroxyvitamin D are at their nadir. Also, nations in the northern hemisphere have borne much of the burden of cases and mortality. In a cross-sectional analysis across Europe, COVID-19 mortality was significantly associated with vitamin D status in different populations [20]. The low mortality rates in Nordic countries are exceptions to the trend towards poorer outcomes in more northerly latitudes, but populations in these countries are relatively vitamin D sufficient owing to widespread supplementation campaigns. Italy and Spain are also exceptions, but prevalence of vitamin D deficiency in these populations is surprisingly common. Additionally, black and minority ethnic people—who are more likely to have vitamin D deficiency because they have darker skin—seem to be worse affected than white people by COVID-19. For example, data from the UK Office for National Statistics shows that black people in England and Wales are more than four times more likely to die from COVID-19 than are white people [21]. In Africa, where they spend more times outdoors and they suffer less from obesity, they seem mostly unaffected by Covid with some of the lowest mortality.

How much of the effect of comorbidities were precipitated by Covid? How much of it is disorganization of the health care system leading to death from other causes? The great variability from state to state indicates that a significant portion is due to disorganization as same pathogen would cause similar patterns in a homogeneous society.

De Larochelambert et al. showed that higher Covid death rates are observed in the [25/65°] latitude and in the [−35/−125°] longitude ranges. The national criteria most associated with death rate are life expectancy and its slowdown, public health context (metabolic and non-communicable diseases (NCD) burden vs. infectious diseases prevalence), economy (growth national product, financial support), and environment (temperature, ultra-violet index). Stringency of the measures settled to fight pandemia, including lockdown, did not appear to be linked with death rate. They concluded that countries that already experienced a stagnation or regression of life expectancy, with high income and NCD rates, had the highest price to pay. This burden was not alleviated by more stringent public decisions [22].

At a time, where epidemic seems to be subsiding in much of the northern hemisphere possibly thanks to immunity having built up naturally and complemented through immunization of most frail, and highly exposed, at a time where recent literature indicates lasting, reactive cross cellular immunity against variants [23][24], given our finding and those of others, some preliminary conclusions can be made. Strict lockdown does not appear to serve a purpose. Neither does completely ignoring the epidemic. Given our awareness that more pandemics will come, it appears that countries would do well to concentrate efforts on rapid location and quarantining of super-spreading cases, improving indoor ventilation so that air does not circulate from room to room in large buildings, improving ventilation in subways and on public transport systems, providing opportunities for spending time outside, increase fitness, reduce obesity, supplement with vitamin D. All such measures would be efficient for droplet and airborne transmitted diseases and would benefit populations’ health by maintaining the strong and strengthening the weak.

## Data Availability

All data is available as public datasets in our references 1 through 11.

## Contributorship Statement

Drs. Sellier and Bricaire conceived the project and assembled the team to conduct the project. Identification of appropriate databases and statistical analysis of data within those databases was conducted by Drs. Cuyugan, Barac, Parvaiz, Bin Jamil, Iqbal, Vally, Koliali, and Sellier. Interpretation of the findings involved all the authors. Dr. Sellier and Dr. Mehl-Madrona wrote the article.

## Data Sharing

The URLs for all datasets used are provided in the text of the article and are listed as references 1 through 11.

## Abbreviations used

SARSCOV-2 = the most recent version of the coronavirus leading to the current pandemic

HVAC: heating, ventilation, and air conditioning
CI: Confidence Interval
INSEE: Institut national de la statistique et des études économiques
**E**UROMOMO: European Mortality Monitoring Project
Statbel: Statistics Belgium
UK: United Kingdom
USA: United States of America
UV: ultraviolet light
GDP: gross domestic product
AC: air conditioning
JRAIA: Japan Refrigeration and Air Conditioning Association
P: probability

